# Impact of a National Immunisation Program on herpes zoster incidence in Australia

**DOI:** 10.1101/2021.08.09.21261775

**Authors:** Jialing Lin, Timothy Dobbins, James G Wood, Carla Bernardo, Nigel P Stocks, Bette Liu

## Abstract

**Objectives:** To evaluate the impact of the National Herpes Zoster (zoster) Immunisation Program in Australia on zoster incidence.

**Methods:** Ecological analysis of zoster incidence related to timing of implementation of the national program in vaccine-targeted (70-79 years) and non-targeted age groups (60-69 and 80-89 years) during January 2013-December 2018 was estimated using interrupted time-series analyses.

**Results:** Prior to program commencement (Jan 2013 to Oct 2016) in patients aged 60-69, 70-79 and 80-89 years, incidence was mostly stable averaging respectively 7.2, 9.6 and 10.8 per 1000 person-years. In the two years following program commencement, incidence fell steadily in those aged 70-79 years, with an estimated decrease of 2.25 (95% CI: 1.34, 3.17) per 1000 person-years per year, with women having a greater decrease than men (2.83 versus 1.68, p-interaction<0.01). In the two non-vaccine-program-targeted age groups there was no evidence of reduction in zoster incidence: 60-69 years, 0.46 (95% CI: -0.46, 1.38) and 80-89 years, 0.11 (95% CI: -1.64, 1.87).

**Conclusions:** Two years after implementation, an estimated 7000 zoster cases were prevented through the national program. With known waning vaccine efficacy, continued surveillance is needed to ensure these early reductions in incidence are sustained.

## Introduction

Herpes zoster (zoster) is a painful rash that occurs following reactivation of the varicella zoster virus. It is more frequently observed in older people and those who are immunocompromised.^1^ It can cause prolonged and severe pain leading to significant health problems including difficulty mobilising and also psychological conditions, such as anxiety, depression, and even social isolation.^2^ The lifetime risk of acquiring zoster is 20-30%, with incidence increasing with age.^3, 4^ A recent study reported annual age-specific zoster incidence of 1.8 per 1000 persons for those aged under 25 years, 6.3 per 1000 for 50-59 year olds, and 19.9 per 1000 for those aged 80 years and over in Australia.^5^

Two effective zoster vaccines are currently registered; one is a live-attenuated vaccine with reported one-dose efficacy of 51.3% against zoster in adults aged 60 years and over^6^ and the other, a recombinant subunit adjuvanted vaccine with two-dose efficacy of 97.2% against zoster in adults aged 50 years and over^7^. In Australia, the live-attenuated vaccine was registered in 2007 but not available until 2013 by private prescription. The recombinant subunit adjuvanted vaccine was registered in 2018 but is not available yet. Since November 2016 in Australia under a National Herpes Zoster Immunisation Program, a single dose of the live-attenuated zoster vaccine has been offered to adults aged 70 years with a time-limited catch-up program for those aged 71-79 years until 2021.^8^ Evaluating the impact of this national program on zoster incidence has been limited by the fact that in Australia zoster is not notifiable in all states and territories,^9^ and the reliability of reporting of zoster to public health units is uncertain with notifications likely to underestimate true rates. Also, internationally there is only one other impact evaluation of a national zoster program.^10^

Therefore, in order to add to the limited knowledge regarding zoster vaccine program effects at a population level, the aim of this study was to evaluate the impact of the National Herpes Zoster Immunisation Program on zoster incidence by using a large national general practice dataset.

## Methods

### Ethics

This study was approved by the MedicineInsight Independent External Data Governance Committee (reference number 2018-015) and the University of New South Wales Human Research Ethics Committee (No. HC180939).

### Data source

MedicineInsight is a large, longitudinal general practice dataset with data extracted from electronic medical records in general practices across Australia. Participating general practices consent to provide de-identified data on an ongoing basis to the MedicineInsight program. The program was developed and is managed by NPS MedicineWise. The dataset contains separate tables with patient demographic records, diagnoses, reasons for encounter, and prescriptions made during patient encounters. Each record is dated and is assigned a unique patient identifier. This enables records related to the same individual to be linked. More details of this dataset are available elsewhere.^11, 12^ In this study, we used an extract containing records from 25% of patients randomly selected from the whole dataset with complete records to 31^st^ December 2018.

### Study population

For this analysis we used presentations recorded in the MedicineInsight dataset from 1^st^ January 2013-31^st^ December 2018 in adults turning 60-89 years old. As people in Australia have unrestricted choice when accessing general practice services, we needed to define a regular attender for a population denominator to estimate incidence. As done previously^13^, we defined our main study population as those who had records of attending the same practice on at least one non-administrative visit each year in the previous two years. Administrative visits were those that only involved administrative notes, emails, faxes, file reviews, phone calls, and phone messages. In addition, patients recorded as deceased in that year, or with missing data on year of birth, sex or zoster diagnosis date were excluded from the study population.

### Definition of herpes zoster cases

For our main analyses we defined a case as having a diagnosis of zoster if they had a term indicating zoster (zoster, HZ, shingles, zona) in either the encounter reason or diagnosis tables but not one of the exclusion terms. Exclusion terms included a word indicating only a possible/suspected diagnosis, or a term indicating the encounter was for vaccination, advice only, or related to a family member (see Appendix Table 1 for full list). Encounters where there was a record of a zoster vaccine administered in the immunisation table on the same date were also excluded as cases. As both the encounter reason and diagnosis tables were free text, to include possible misspellings, we used the SPEDIS function with cost ≤30 in SAS to identify closely related spellings to our zoster terms.

### Data analysis

All records that met the case definition with the same patient identifier and record date were considered duplicates and removed before data analysis. If participants had more than one record of a zoster diagnosis over the study period, records that were separated by more than 180 days during the observation period were considered as distinct episodes of zoster^14^ and contributed to counts of events. For each calendar year of data, we divided participants into three groups based on the age they turned in that year: 60-69, 70-79 (targeted for vaccination), and 80-89 years old. Zoster incidence in each month and the year of observation was estimated by calculating the number of zoster episodes recorded in that period divided by person-years of the study population meeting the regular attender definition in that year.

For estimating annual zoster incidence before and after the national program, we separated data into the three-year time period from November 2013 to October 2016 which was considered “before the program”, and the two-year period from November 2016 to October 2018 which was considered “after the program”. We estimated zoster incidence in each period (before and after the program) and the relative and absolute reductions in incidence comparing the periods, stratified by age and sex. We also calculated incidence before and after the program in those 70-79 years by practice jurisdiction, remoteness status^15^ and area-level socioeconomic disadvantage using the index of relative socioeconomic advantage and disadvantage (IRSAD)^16^.

We used an interrupted time-series model^17^ to further investigate the effect of the national program on zoster incidence. We defined January 2013-October 2016 as the pre-program period and November 2016-December 2018 as the post-program period. A segmented regression was used, including terms to estimate the base level of incidence at the beginning of the series, the pre-program trend in incidence, the change in level of incidence immediately after the program and the change in trend of incidence after the program. We assessed serial autocorrelation graphically and via the Durbin-Watson statistic, accounting for autocorrelation where necessary by including an auto-regressive term. Model assumptions were checked using graphical summaries of residuals. In addition, we calculated the total number of zoster cases averted in the vaccine-targeted age group in Australia by applying our estimates of absolute reduction to population estimates from the 2016 census.^18^

All analyses were conducted with SAS v. 9.4 (SAS Institute Inc. Cary, North Carolina, USA).

### Sensitivity analysis

In the first sensitivity analysis we re-defined the study population, to be consistent with the Royal Australian College of General Practitioners (RACGP)^19^ which defines a regular attender as a patient who attended the same practice for at least three non-administrative visits on different days within the last two years. For the second, we broadened the definition of a zoster case by including those with a prescription record of a zoster-specific antiviral (see Appendix Table 2 for details). This included acyclovir (800 mg) with 35 tablets, valacyclovir (500 mg) with 42 tablets, or famciclovir (250 mg) with 21 tablets.^20^

## Results

For each analysis year, there were: between 50814 and 62677 regular attenders aged 70-79 years, 82248 to 89542 aged 60-69 years, and 27166 to 29866 aged 80-89 years. For the vaccine-targeted age group (70-79 years), there were 2439 patients who met the zoster case definition over the five-year period, Nov 2013-Oct 2018. For the two non-vaccine-targeted age groups, 60-69 and 80-89 years of age, the corresponding figures were 1495 and 550, respectively.

Table 1 shows the crude average annual incidence of zoster before and after program commencement. Zoster incidence per 1000 person-years before and after the program was 9.5 versus 7.5 for those 70-79 years, 7.7 versus 8.3 for those 60-69 years and 9.0 versus 9.3 for those 80-89 years. Among those aged 70-79 years the absolute reduction in incidence was 2.0 per 1000 person-years or a relative reduction of 21%. Such falls were not observed in either the 60-69 or the 80-89 years age groups. Women had consistently higher incidence than men in all age groups both before and after program commencement and falls in incidence were observed for both men and women aged 70-79 years. Among the vaccine-targeted population (aged 70-79 years) we also found incidence decreased after the program in all remoteness and socioeconomic status categories, and in most jurisdictions, except the Northern Territory and Western Australia (Appendix Table 3).

**Table 1.** Crude annual incidence of herpes zoster per 1000 person-years before and after implementation of the National Herpes Zoster Immunisation Program among patients attending General Practices in Australia, by age and sex.

| Age, sex | Before program<br>(Nov 2013-Oct 2016) | After program<br>(Nov 2016-Oct 2018) |
| --- | --- | --- |
| **60-69 y** |  |  |
| Overall | 7.7 (1979/256157) | 8.3 (1495/179146) |
| Men | 6.3 (753/119315) | 6.6 (547/82576) |
| Women | 9.0 (1226/136842) | 9.8 (948/96570) |
| **70-79 y** |  |  |
| Overall | 9.5 (1518/160727) | 7.5 (921/122779) |
| Men | 8.4 (626/74847) | 6.6 (381/57692) |
| Women | 10.4 (892/85880) | 8.3 (540/65087) |
| **80-89 y** |  |  |
| Overall | 9.0 (756/83874) | 9.3 (550/59234) |
| Men | 7.7 (272/35392) | 8.9 (224/25130) |
| Women | 10.0 (484/48482) | 9.6 (326/34104) |

Incidence is shown by the incidence rate (total number of zoster episodes/total number of person-years).

Trends in zoster incidence pre- and post-program by age group are shown in Figure 1. Prior to program commencement in patients aged 60-69, 70-79 and 80-89 years, incidence was mostly stable averaging respectively 7.2, 9.6 and 10.8 per 1000 person-years. However, following the program commencement, the incidence fell steadily in those aged 70-79 years.

**Fig. 1.**
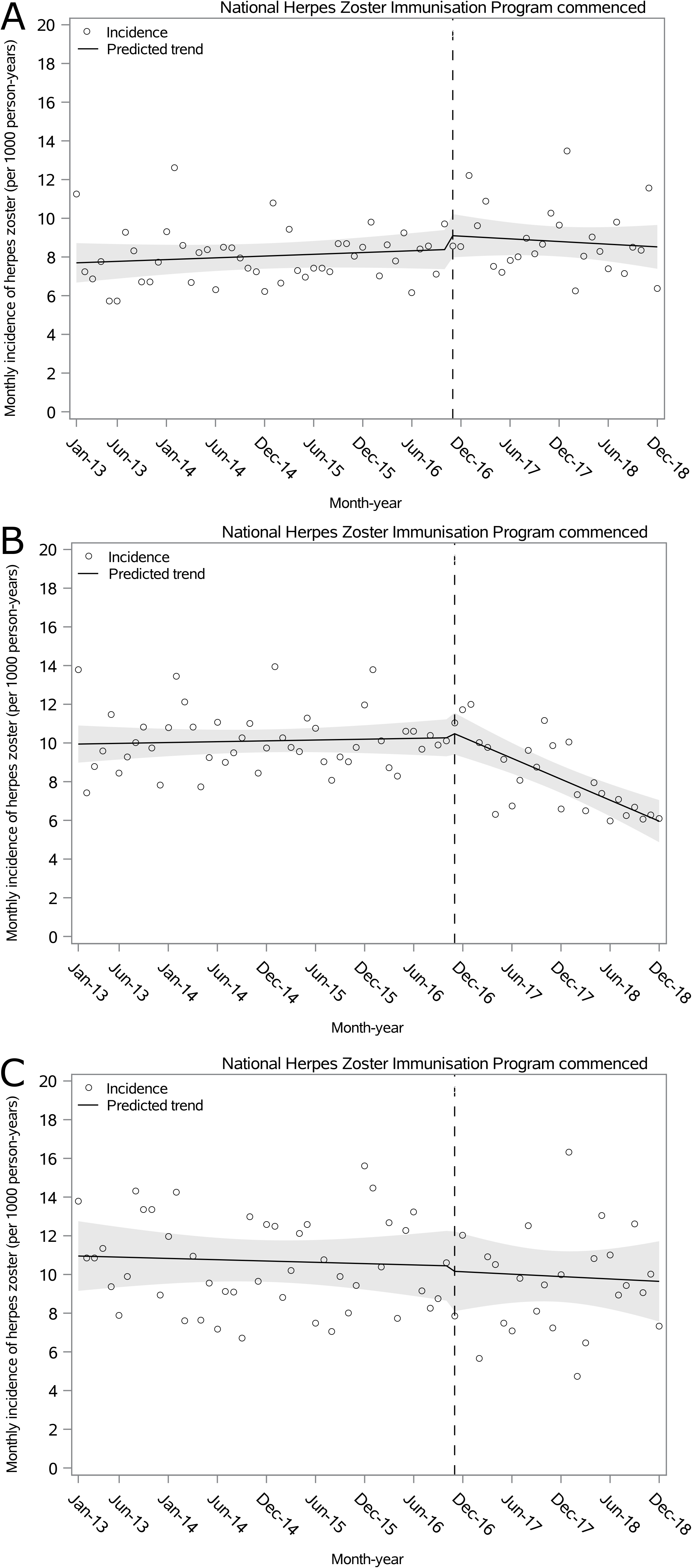
Monthly incidence of herpes zoster per 1000 person-years among patients aged (A) 60-69, (B) 70-79 and (C) 80-89 years old attending General Practices during Jan 2013-Dec 2018 in Australia.

From the interrupted time-series analysis, the trend in zoster incidence in the vaccine-targeted age group decreased by 2.25 (95% CI: 1.34, 3.17) per 1000 person-years per year (Table 2). Women had a greater decrease than men (2.83 versus 1.68, test for interaction <0.01). By contrast, there was no evidence of reductions in zoster incidence trends for the two non-vaccine-targeted age groups.

**Table 2.**
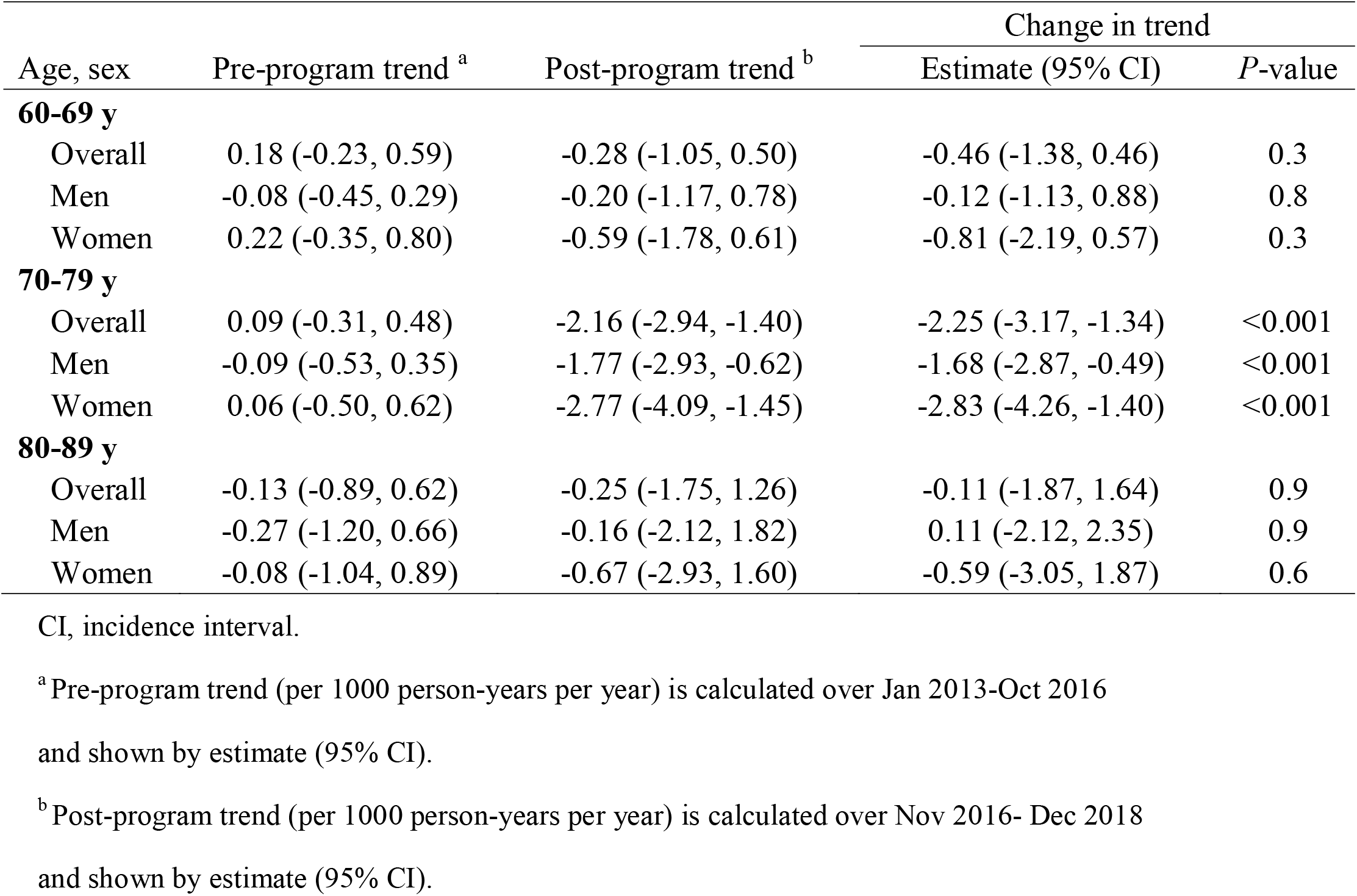
Impact of the National Herpes Zoster Immunisation Program on trends in incidence of herpes zoster among patients attending General Practices in Australia, by age and sex.

| Age, sex | Pre-program trend <sup>a</sup> | Post-program trend <sup>b</sup> | Change in trend |  |
| --- | --- | --- | --- | --- |
|  |  |  | Estimate (95% CI) | *P*-value |
| **60-69 y** |  |  |  |  |
| Overall | 0.18 (-0.23, 0.59) | -0.28 (-1.05, 0.50) | -0.46 (-1.38, 0.46) | 0.3 |
| Men | -0.08 (-0.45, 0.29) | -0.20 (-1.17, 0.78) | -0.12 (-1.13, 0.88) | 0.8 |
| Women | 0.22 (-0.35, 0.80) | -0.59 (-1.78, 0.61) | -0.81 (-2.19, 0.57) | 0.3 |
| **70-79 y** |  |  |  |  |
| Overall | 0.09 (-0.31, 0.48) | -2.16 (-2.94, -1.40) | -2.25 (-3.17, -1.34) | <0.001 |
| Men | -0.09 (-0.53, 0.35) | -1.77 (-2.93, -0.62) | -1.68 (-2.87, -0.49) | <0.001 |
| Women | 0.06 (-0.50, 0.62) | -2.77 (-4.09, -1.45) | -2.83 (-4.26, -1.40) | <0.001 |
| **80-89 y** |  |  |  |  |
| Overall | -0.13 (-0.89, 0.62) | -0.25 (-1.75, 1.26) | -0.11 (-1.87, 1.64) | 0.9 |
| Men | -0.27 (-1.20, 0.66) | -0.16 (-2.12, 1.82) | 0.11 (-2.12, 2.35) | 0.9 |
| Women | -0.08 (-1.04, 0.89) | -0.67 (-2.93, 1.60) | -0.59 (-3.05, 1.87) | 0.6 |

CI, incidence interval.
<sup>a</sup> Pre-program trend (per 1000 person-years per year) is calculated over Jan 2013-Oct 2016 and shown by estimate (95% CI).
<sup>b</sup> Post-program trend (per 1000 person-years per year) is calculated over Nov 2016- Dec 2018 and shown by estimate (95% CI).

### Sensitivity analyses

Use of the RACGP definition of a regular attender did not change the patterns in incidence. In those aged 70-79 years the post-program decrease in incidence was 1.98 (95% CI: 1.13, 2.84) per 1000 person-years per year whilst there was no change in those 60-69 and those 80-89 years (see Appendix Table 4).

Using the broader zoster case definition resulted in higher incidences but the patterns were similar. Before and after the program for those aged 60-69, 70-79, 80-89 years respectively: 9.4 versus 10.0, 11.5 versus 9.1, 11.0 versus 11.2 per 1000 person-years. There was also evidence of a reduction in incidence trends in the 70-79 years age group (reduction: 2.45 (95% CI: 1.63, 3.27) per 1000 person-years per year) but not in either the 60-69 or the 80-89 years age groups (see Appendix Table 5).

## Discussion

To the best of our knowledge, this is the first study to estimate the impact of the National Zoster Immunisation Program on zoster incidence in Australia. We found that in the two years after implementation, zoster incidence decreased by 2.25 per 1000 person-years per year or by about 40% from pre-program levels in the vaccine-targeted population (those aged 70-79 years); no significant falls were recorded in those aged 60-69 and 80-89 years.

Our findings are consistent with studies in other countries and regions that have also implemented and evaluated zoster immunisation programs with live-attenuated zoster vaccines. England introduced a zoster immunisation program for 70 year olds with phased catch-up for those 71-79 years in September 2013.^10^ After three years, vaccine coverage was reported as 60% in the vaccine-targeted population, and zoster incidence was reported to decrease by 3.1 per 1000 person-years. In the Canadian province of Ontario, a zoster immunisation program was introduced in September 2016 for adults aged 65-70 years.^21^ Results of a national survey showed vaccine uptake was 21% among participants aged ≥50 years in 2016^22^ and the monthly incidence of medically-attended zoster in the 2 years after program implementation was reported to decrease by 1 per 10,000 population^21^.

Some of the differences in the estimated impact of the zoster immunisation program in Australia compared to other regions could be due to differences in zoster case ascertainment used in this study and vaccine coverage achieved. Estimates of zoster vaccine uptake in Australia are limited. A report based on data from the Australian Immunisation Register suggested that cumulative vaccine coverage 23 months after program commencement in Australia was 31.2% among adults aged 70-79 years but this is considered an underestimate due to underreporting.^23^ Using this same general practice database (MedicineInsight) in a separate study we have estimated higher coverage (47%) in the targeted population 2 years after program commencement.^13^

Women had both higher incidence of zoster but also greater reductions in incidence than men following program implementation, and this might be explained by higher vaccine uptake in women than men.^13, 23^ This sex difference was also observed in the Canadian study.^21^ The fall in incidence post-program was also observed to be relatively uniform across Australian regions (by remoteness) and socioeconomic groups, and also in most jurisdictions, although not in the Northern Territory and Western Australia. The lack of a reduction in incidence in the Northern Territory may be due to the small number of participating practices in our sample and therefore a lack of power to detect a difference; although vaccine coverage in the Northern Territory is also reported to be substantially lower^23^. The lack of change in incidence detected in Western Australia is more difficult to explain as coverage is reportedly similar to other jurisdictions^23^ and further investigation is required to confirm this difference.

Study strengths include the use of a large national general practice dataset of electronic medical records allowing us sufficient numbers of cases in age groups to investigate changes over time. However, there are some limitations. Firstly, we only studied people attending GPs so zoster cases diagnosed in settings such as emergency departments or hospitals or at GPs not participating in the MedicineInsight database would be missed but this proportion is likely to be small and unlikely to differ between the pre- and post-program periods examined. Secondly, there is the potential for misclassification of zoster cases and we were unable to validate the case records. However, we used the same case definition consistently over time and in different age groups and we included a range of terms and misspellings to improve detection and exclude questionable cases. The fact we observed a fall in incidence coinciding with the time of program implementation and only in vaccine-targeted age groups suggest that our definition was valid.

## Conclusion

In summary, our study shows that the recently implemented National Herpes Zoster Immunisation Program in Australia has had a significant impact on reducing zoster incidence among the vaccine-targeted population and that the level of reduction appears to be consistent with other regions in the world that have similar programs in place, given differences in vaccine uptake. Based on Australian census data^24^ for the population aged 70-79 years (N=1,540,376), we estimate about 7000 zoster cases would have been averted two years after the program commenced. Given the known waning of vaccine efficacy for the live-attenuated vaccine,^25^ longer-term evaluation of this program is also needed to ensure that zoster incidence continues to decrease.

## Data Availability

The provision of information to third parties with an interest in MedicineInsight data is subject to a rigorous and formal approval process and is guided by the MedicineInsight independent external Data Governance Committee. This Committee includes GPs, consumer advocates, privacy experts, and researchers.

## Acknowledgements

We thank the NPS MedicineWise MedicineInsight for providing the data for this study. J. Lin is supported by the University International Postgraduate Awards under a UNSW Tuition Fee Scholarship and a School of Population Health Stipend Scholarship. B. Liu is supported by an NHMRC Fellowship.

