## Appendix for "Impact of a National Immunisation Program on herpes zoster incidence in Australia"

**Table 1** Exclusion terms for a diagnosis of herpes zoster.

Terms

“?”, “possible”, “probable”, “presume”, “suspected”, “doubt”, “consider”, “like”, “assume”, “maybe”, “screen”, “worried”, “varicella”, “neuralgia”, “in case of”, “zostervax”, “zostavax”, “zostavac”, “zostervac”, “zostervacs”, “zostervacx”, “zosterwax”, “zosteravax”, “zostax”, “zostovax”, “zostavzax”, “zostarzax”, “zostavacx”, “zostrix”, “zosterix”, “shingles vax”, “vaccine”, “vaccination”, “immunisation”, “prevent”, “prevention”, “needle”, “shot”, “ZVL”, “immunity”, “igg”, “test negative”, “previous”, “exposure”, “past”, “recall”, “invitation”, “email”, “phone”, “follow”, “sms”, “message”, “review”, ”discuss”, “consult”, “letter”, “family history”, ”wife”, “mother”, “husband”, “son”, “sister”, “contact”, “exclude”, “not”, “no”

**Table 2** Formulation of herpes zoster-specific antiviral treatment.

| Antiviral treatment | Medication name | Strength | Quantity | Form |
| --- | --- | --- | --- | --- |
| Acyclovir | “acyclovir”, aciclovir”, “acycloguanosine”, “aciclovirum”, “acv”, “zovirax” | “800mg” | 35 | “tablet” |
| Valacyclovir | “valacyclovir”, “valaciclovir”, “valtrex”, “valacyclovirum” | “500mg” | 42 | “tablet” |
| Famciclovir | “famciclovir”, “famciclovirum”, “famvir”, “fcv” | “250mg” | 21 | “tablet” |

**Table 3** Crude annual incidence of herpes zoster per 1000 person-years before and after implementation of the National Herpes Zoster Immunisation Program among 70-79 years old patients attending General Practices in Australia, by practice jurisdiction, remoteness and socioeconomic status.

| Characteristics | Pre-program  (Nov 2013-Oct 2016) | Post-program  (Nov 2016-Oct 2018) |
| --- | --- | --- |
| Jurisdiction |  |  |
| ACT | 11.4 (32/2803) | 10.0 (21/2103) |
| NSW | 9.5 (558/58894) | 7.5 (340/45165) |
| NT | 1.3 (<5/1498) | 5.0 (6/1194) |
| QLD | 10.7 (248/23282) | 7.5 (137/18377) |
| SA | 8.9 (45/5039) | 6.6 (26/3929) |
| TAS | 10.6 (160/15130) | 8.7 (101/11626) |
| VIC | 9.6 (335/34851) | 6.8 (173/25418) |
| WA | 7.2 (138/19230) | 7.8 (117/14967) |
| Remoteness status |  |  |
| Major cities | 9.7 (896/92060) | 7.5 (531/70800) |
| Inner regional | 9.5 (431/45431) | 7.9 (272/34378) |
| Outer regional/remote/ very remote | 8.2 (191/23236) | 6.7 (118/17601) |
| Quintile of socioeconomic disadvantage ^*^ |  |  |
| 1 (most disadvantaged) | 8.8 (290/33014) | 6.4 (159/24663) |
| 2 | 9.2 (248/26859) | 7.4 (151/20351) |
| 3 | 9.0 (357/39858) | 8.1 (251/30878) |
| 4 | 10.1 (240/23705) | 8.1 (152/18813) |
| 5 (most advantaged) | 10.3 (377/36629) | 7.3 (201/27591) |

Incidence is shown by the incidence rate (total number of zoster episodes/total number of person-years).

ACT, Australian Capital Territory; NSW, New South Wales; NT, Northern Territory; QLD, Queensland; SA, South Australia; TAS, Tasmania; VIC, Victoria; WA, Western Australia.

^*^ Data not shown for N=1145 patients who had missing data on quintile of socioeconomic disadvantage.

**Table 4** Impact of the National Herpes Zoster Immunisation Program on trends in incidence of herpes zoster among RACGP patients attending General Practices in Australia, by age and sex.

| Age, sex | Pre-program trend ^a^ | Post-program trend ^b^ | Change in trend | |
| --- | --- | --- | --- | --- |
|  |  |  | Estimate (95% CI) | *P*-value |
| **60-69 y** |  |  |  |  |
| Overall | 0.10 (-0.27, 0.46) | -0.52 (-1.20, 0.16) | -0.62 (-1.43, 0.20) | 0.1 |
| Men | -0.12 (-0.57, 0.34) | -0.30 (-1.26, 0.68) | -0.18 (-1.29, 0.93) | 0.8 |
| Women | 0.17 (-0.34, 0.69) | -0.93 (-1.97, 0.12) | -1.10 (-2.31, 0.11) | 0.08 |
| **70-79 y** |  |  |  |  |
| Overall | 0.01 (-0.36, 0.38) | -1.97 (-2.70, -1.26) | -1.98 (-2.84, -1.13) | <0.001 |
| Men | -0.19 (-0.64, 0.26) | -2.03 (-3.23, -0.82) | -1.84 (-3.08, -0.60) | <0.001 |
| Women | -0.11 (-0.52, 0.31) | -2.42 (-3.42, -1.42) | -2.31 (-3.38, -1.24) | <0.001 |
| **80-89 y** |  |  |  |  |
| Overall | -0.15 (-0.81, 0.50) | -0.52 (-1.82, 0.78) | -0.37 (-1.89, 1.15) | 0.6 |
| Men | -0.26 (-0.91, 0.39) | -1.09 (-2.64, 0.46) | -0.83 (-2.50, 0.84) | 0.3 |
| Women | -0.04 (-0.85, 0.78) | -0.61 (-2.53, 1.32) | -0.57 (-2.66, 1.52) | 0.6 |

RACGP, Royal Australian College of General Practitioners, defines a regular attender as a patient who attended the same practice for at least three non-administrative visits on different days within the last two years; CI, incidence interval.

^a^ Pre-program trend (per 1000 person-years per year) is calculated over Jan 2013-Oct 2016 and shown by estimate (95% CI).

^b^ Post-program trend (per 1000 person-years per year) is calculated over Nov 2016- Dec 2018 and shown by estimate (95% CI).

**Table 5** Impact of the National Herpes Zoster Immunisation Program on trends in incidence of herpes zoster with broader definition ^*^ among patients attending General Practices in Australia, by age and sex.

| Age, sex | Pre-program trend ^a^ | Post-program trend ^b^ | Change in trend | |
| --- | --- | --- | --- | --- |
|  |  |  | Estimate (95% CI) | *P*-value |
| **60-69 y** |  |  |  |  |
| Overall | 0.18 (-0.24, 0.60) | -0.13 (-0.95, 0.70) | -0.31 (-1.28, 0.67) | 0.5 |
| Men | 0.01 (-0.37, 0.39) | 0.23 (-0.79, 1.25) | 0.22 (-0.37, 0.39) | 0.7 |
| Women | 0.22 (-0.24, 0.68) | -0.33 (-1.49, 0.82) | -0.55 (-1.76, 0.66) | 0.4 |
| **70-79 y** |  |  |  |  |
| Overall | 0.07 (-0.24, 0.38) | -2.38 (-3.16, -1.59) | -2.45 (-3.27, -1.63) | <0.001 |
| Men | -0.04 (-0.78, 0.69) | -1.97 (-3.68, -0.26) | -1.93 (-3.80, -0.06) | 0.047 |
| Women | 0.23 (-0.28, 0.73) | -2.65 (-3.91, -1.39) | -2.88 (-4.20, -1.55) | <0.001 |
| **80-89 y** |  |  |  |  |
| Overall | -0.05 (-0.89, 0.78) | -0.78 (-2.51, 0.96) | -0.73 (-2.71, 1.26) | 0.5 |
| Men | -0.20 (-1.20, 0.80) | -0.36 (-2.47, 1.75) | -0.16 (-2.57, 2.25) | 0.9 |
| Women | 0.05 (-1.00, 1.09) | -1.30 (-3.76, 1.16) | -1.35 (-4.02, 1.33) | 0.3 |

CI, incidence interval.

^*^ Defined zoster cases as a diagnosis of zoster in either the encounter reason or diagnosis tables or a receipt of specific zoster treatment in the prescription table.

^a^ Pre-program trend (per 1000 person-years per year) is calculated over Jan 2013-Oct 2016 and shown by estimate (95% CI).

^b^ Post-program trend (per 1000 person-years per year) is calculated over Nov 2016-Dec 2018 and shown by estimate (95% CI).
